# Severity of Omicron (B.1.1.529) and Delta (B.1.617.2) SARS-CoV-2 infection among hospitalised adults: a prospective cohort study in Bristol, United Kingdom

**DOI:** 10.1101/2022.06.29.22277044

**Authors:** Catherine Hyams, Robert Challen, Robin Marlow, Jennifer Nguyen, Elizabeth Begier, Jo Southern, Jade King, Anna Morley, Jane Kinney, Madeleine Clout, Jennifer Oliver, Sharon Gray, Gillian Ellsbury, Nick Maskell, Luis Jodar, Bradford Gessner, John McLaughlin, Leon Danon, Adam Finn, The Avon CAP Research Group

## Abstract

**Background:** There is an urgent public health need to evaluate disease severity in adults hospitalised with Delta and Omicron SARS-CoV-2 variant infections. However, limited data exist assessing severity of disease in adults hospitalised with Omicron SARS-CoV-2 infections, and to what extent patient-factors, including vaccination, age, frailty and pre-existing disease, affect variant-dependent disease severity.

**Methods:** A prospective cohort study of adults (≥18 years of age) hospitalised with acute lower respiratory tract disease at acute care hospitals in Bristol, UK conducted over 10-months. Delta or Omicron SARS-CoV-2 infection was defined by positive SARS-CoV-2 PCR and variant identification or inferred by dominant circulating variant. We constructed adjusted regression analyses to assess disease severity using three different measures: FiO_2_ >28% (fraction inspired oxygen), World Health Organization (WHO) outcome score >5 (assessing need for ventilatory support), and hospital length of stay (LOS) >3 days following admission for Omicron or Delta infection.

**Findings:** Independent of other variables, including vaccination, Omicron variant infection in hospitalised adults was associated with lower severity than Delta. Risk reductions were 58%, 67%, and 16% for supplementary oxygen with >28% FiO_2_ [Relative Risk (RR)=0·42 (95%CI: 0·34-0·52), *P*<0.001], WHO outcome score >5 [RR=0·33 (95%CI: 0·21-0·50), *P*<0.001], and to have had a LOS>3 days [RR=0·84 (95%CI: 0·76-0·92), *P*<0.001]. Younger age and vaccination with two or three doses were also independently associated with lower COVID-19 severity.

**Interpretation:** We provide reassuring evidence that Omicron infection results in less serious adverse outcomes than Delta in hospitalised patients. Despite lower severity relative to Delta, Omicron infection still resulted in substantial patient and public health burden and an increased admission rate of older patients with Omicron which counteracts some of the benefit arising from less severe disease.

**Funding:** AvonCAP is an investigator-led project funded under a collaborative agreement by Pfizer.

**RESEARCH IN CONTEXT:** *Evidence before this study:* The burden of COVID-19 on hospital services is determined by the prevalence and severity of SARS-CoV-2 variants, and modified by individual factors such as age, frailty and vaccination status. Real world data suggest that vaccine effectiveness is lower and may wane faster over time against symptomatic disease with Omicron (B.1.1.529) than with Delta (B.1.617.2) SARS-CoV-2 variant. However, numbers of hospitalisations as a case proportion during the Omicron wave have been considerably lower than previous waves. Several reports have compared the risk of hospitalisation or severe disease based on SARS-CoV-2 variant, some suggesting that Omicron is probably less severe than Delta in vaccinated and unvaccinated individuals.

*Added value of this study:* This study provides robust data assessing the relative severity of Delta and Omicron SARS-CoV-2 variants in patients admitted to hospital, including the first analysis assessing risk for any positive pressure ventilatory support, as well as risk of supplementary oxygen requirement and extended hospital admission, that may guide resource planning in hospitals. We found evidence that infection with Omicron was associated with a milder clinical course following hospital admission than that caused by Delta and that vaccination was independently associated with lower in-hospital disease severity using these three separate severity measures. Specifically, compared to Delta, Omicron-related hospitalizations were 58%, 67%, and 16% less likely to require high flow oxygen >28% FiO_2_, positive pressure ventilatory support or more critical care, and to have a hospital stay lasting more than three days, respectively. This study reports the considerable morbidity resulting from Omicron infection, with 18% of Omicron admissions requiring oxygen supplementation FiO_2_ >28%, 6% requiring positive pressure ventilation, 62% needing hospitalization ≥four days, and 4% in-hospital mortality. In determining the reduced requirement of increased oxygen requirement and total positive pressure requirement, including non-invasive ventilation, this analysis should contribute to future hospital care and service planning assessments.

*Implications of all the available evidence:* The risk of severe outcomes following SARS-CoV-2 infection is substantially lower for Omicron than for Delta, with greater reductions for more severe disease outcomes. Significant variation in risk occurs with age and vaccination status, with older and unvaccinated individuals remaining at particular risk of adverse outcome. These results highlight the importance of maintaining high levels of vaccine coverage in patient groups at risk of severe disease. The impact of lower severity Omicron-related hospitalization must be balanced against increased transmissibility and overall higher numbers of infections with this variant and there remains a substantial patient and public health burden. The increased admission rate of older patients with Omicron counteracts some of the benefit arising from less severe disease. Despite the risk reduction in high level oxygen supplementation requirement and high dependency care with Omicron compared to earlier variants at the individual level, healthcare systems could still be overwhelmed.

## INTRODUCTION

The emergence of SARS-CoV-2 has resulted in a global pandemic, with multiple variants detected since the wild-type virus emerged. The Delta variant (B.1.617.2) was first detected in India in March 2021, spread rapidly [1] and resulted in a sharp rise in SARS-CoV-2 cases in the United Kingdom [2]. The Omicron (B.1.1.529) variant, first detected in South Africa in November 2021, rapidly replaced Delta as the main circulating variant globally and resulted in a fourth wave of SARS-CoV-2 infection in the UK. By 26^th^ December 2021, 95% of UK SARS-CoV-2 cases were estimated to be caused by Omicron [3].

SARS-CoV-2 variants may differ in their capacity to infect and transmit, susceptibility either to vaccine- or infection-derived immunity and clinical phenotype including disease severity. Laboratory data suggest that vaccine-induced neutralizing antibody activity is lower against Delta [4,5] and Omicron variants [6,7] than against the Alpha variant and wild/prototype viral strains. Real world data subsequently confirmed vaccine effectiveness is lower against symptomatic disease with Omicron than with Delta and other prior variants [8,9], with protection that may also wane faster over time [10-13]. However, numbers of hospitalisations as a case proportion during the Omicron wave have been considerably lower than previous waves [2,14]. Observational data suggest that vaccines remain effective against Omicron-related hospitalization [15,16].

Several reports [11,17-19] have compared the risk of hospitalization or severe disease based on SARS-CoV-2 variant, some suggesting that Omicron is probably less severe than Delta in vaccinated and unvaccinated individuals [11,17,18]. However, a recent preliminary report found no difference in severity between Omicron and Delta [19]. While some studies have stratified results by vaccination status [11,17,18] and age [17], none have comprehensively evaluated the simultaneous impact of these and other important factors (e.g., chronic medical conditions, frailty) on more detailed measures of relative clinical severity in UK hospitalized patients.

In this study, we assessed the relative impact of SARS-CoV-2 variant on three separate COVID-19-related severity measures in adults hospitalised with SARS-CoV-2 infection, after adjusting for important potential confounders, including age, frailty, and vaccination status.

## METHODS

### Study design and study population

We are conducting a prospective cohort study of adults hospitalised with acute respiratory illness to North Bristol and University Hospitals Bristol and Weston NHS Trusts and now report those presenting 1^st^ June 2021 - 28^th^ March 2022 inclusive. All adults aged ≥18 years admitted to these hospitals were screened for respiratory disease signs/symptoms [20]. Of these, patients who tested positive for COVID-19 (detailed below) were included. Details of the UK COVID-19 Vaccination Programme are provided in Supplementary Data 1.

### Data Collection

Clinical data were collected from electronic and paper patient records and recorded on an electronic case report form using REDCap software [21]. Variables collected included: gender; age in years; ethnicity; patient pre-existing diseases; vaccination status; age (categorical); markers of frailty (Charlson Comorbidity Index [CCI] [22], QCovid2 hazard, [23] Rockwood score [24]); immunosuppressive therapy; residence in a care home; index of multiple deprivation (IMD); symptom duration prior to admission; hospital site and study week number. We also collected data about treatment requirement, including: hospital length of stay (LOS); need for supplementary oxygen and level of oxygen provided; requirement for positive pressure ventilatory support.

Data collection was undertaken by individuals not involved in data analysis and collection of data methods were identical for patients with Omicron and Delta variant infections. Vaccination records for each study participant were obtained by team members unaware of subjects’ SARS-CoV-2 test results from linked hospital and GP records, including details of vaccination brand and administration date. The QCovid2 hazard score was calculated for each admission [23], estimating risk of hospitalisation or death due to COVID-19 infection. We also determined the CCI score [22,25], with higher scores indicating not only a greater mortality risk but also more severe comorbid conditions, and the Rockwood frailty score[24], a 9-point scale with score ≥5 indicating frailty.

### Case definitions and exclusions

SARS-CoV-2 infection was defined as respiratory disease and a positive test result for SARS-CoV-2 on/during hospitalisation or within 7 days prior to hospital admission, using the established UK Health Security Agency (UKHSA) diagnostic assay deployed at the time. Infection with a particular SARS-CoV-2 variant was likewise determined using the standard UKHSA variant identification methods deployed at the time. Initially, Delta was identified by S-gene positive PCR (SGTP), which was replaced on 11^th^ May 2021 by the P681R target following validation. Omicron was identified as S-gene negative (SGNP) and then by K417N mutation [26]. Cases without variant identification results admitted between the 1^st^ June 2021 and 7^th^ November 2021 were inferred to be Delta, and those after 7^th^ Feb 2022 were inferred to be Omicron. These two dates represent the first date of a proven Omicron admission and the last date of a proven Delta admission in the Bristol area, respectively.

Patients developing symptoms >10 days prior to hospitalisation were excluded to avoid including individuals who may have falsely tested negative for SARS-CoV-2 upon admission because they presented to hospital late in their clinical course as were individuals who had been inpatients for >28 days prior to their first positive SARS-CoV-2 PCR test. Patients who were partially vaccinated, i.e. had received only one dose or a second dose <7 days before the time of symptom onset, and those who had previously recovered from COVID-19 and had not been vaccinated, were also excluded from the analysis, as their immunological status was uncertain and would prevent meaningful comparison between vaccinated and unvaccinated individuals.

Individuals were defined as unvaccinated if they had never received any COVID-19 vaccine. Immunisation with only two vaccine doses was defined as receipt of two doses with ≥7 days elapsing between symptom onset and the second dose (and no third dose). Immunisation with three doses was defined as receipt of a third vaccine dose after receiving two doses, with ≥7 days elapsing between the third dose and symptom onset. Only patients being admitted with COVID-19 for the first time were included, as repeat admissions may be expected to follow different clinical courses.

### Outcome and covariates

The primary comparison is between cases infected with SARS-CoV-2 Delta variant and SARS-CoV-2 Omicron variant in hospitalised adults. To investigate disease severity and consider the hypothesis that Omicron is associated with less severe outcomes in patients hospitalised with COVID-19, we considered three binary outcomes measured on the seventh day following admission: (1) maximum oxygen requirement FiO_2_ >28%, (2) a WHO outcome score greater than 5 (i.e. ventilation or intubation required or death) within the first seven days of hospitalisation (which also implies a maximum oxygen requirement FiO_2_ > 28%) [27], and (3) length of stay longer than three days. The WHO outcome score, or clinical progression scale is based on the level of invasive ventilation required and, in admitted patients, scores range from 4 to 10 (4 = room air, 5 = oxygen supplementation required, 6 = non-invasive positive pressure ventilation [NIPPV] usage, and 7-9 needing endotracheal intubation). A score of 10 represents death. Outcome measures were chosen to reflect different operational concerns for hospitals, in particular the adequacy of oxygen supply, availability of high dependency care, and a measure of overall hospital bed utilisation. We measured the scores at day seven to include relevant data from the hospital admission and to prevent complications with right censoring. Thresholds for the outcomes used here were determined empirically following an initial investigation, and a sensitivity analysis conducted on the level of each threshold. Charlson Comorbidity Index and age were divided into categories. For CCI the scores were categorised into none (CCI of 0), mild (1-2), moderate (3-4) or severe (5 or more) and have an approximately even spilt across the data. For age, bands of 18-34years (y), 35-49y, 50-64y, 65-74y, 75-84y and 85y and over, were chosen as approximately even categories with more resolution in older age groups.

### Statistical Analysis

The demographics of patients with Delta and Omicron infection were described using mean ± standard deviation (SD) or median (interquartile range [IQR]) and compared using Fisher exact tests for categorical variables, two sided Kolmogorov–Smirnov tests for continuous variables and two-sided Wilcoxon-Mann-Whitney tests for score variables. Where categorical data were missing, this comparison was performed with both missing data removed and with missing data included as a separate category (Table 1). There were no missing data items for continuous variables. We examined trends in patients hospitalised over the study period, determining the proportion of hospitalisations with each variant, by vaccination status (unvaccinated, two or three dose of any COVID-19 vaccine) and age group.

The complex interaction between the dynamics of UK variant distribution and the progressing age-stratified booster campaign rollout was hypothesised to influence both the risk profile of patients hospitalised and severity of outcome once hospitalised. The outcomes are relatively frequent in both groups which can make interpretation of odds ratios resulting from logistic regression complex [28]. Thus, we used Poisson regression with a robust error variance [28] to estimate the relative risk of each of the three outcomes.

The primary comparison was the impact of the variant on hospital outcomes. Other covariates were identified that might also influence, with variable distribution by variant provided (Supplementary Table 3). Covariates were screened for systematic biases due to missing data, resulting in removal of IMD (Supplementary Tables 4, 5). Remaining missing data were imputed using the fully conditional specification approach implemented using the ‘mice‘ package in R [29]. Multiple imputations were performed, with full analysis repeated for each imputation. The resulting coefficients for each imputation were combined using a mixture distribution, assuming normally distributed beta coefficients. For each factor and outcome, the univariate relative risk was estimated, followed by a multivariable model of a linear combination of genomic variant, vaccination, age and CCI [22]. As a sensitivity analysis these were fully adjusted using the remaining covariates, gender, ethnicity, immunosuppressive therapy, residence in a care home, duration of symptoms prior to admission, hospital site, and study week number. Additional sensitivity analyses were conducted using different combinations of age and frailty markers, including CCI as a continuous variable, Rockwood score [24] and QCovid2 hazard rate (which includes age) [23]. Further sensitivity analyses were conducted looking at different threshold values of each of the three binary outcomes.

Statistical analyses were performed in R, version 4.0.2 [30]. Robust Poisson regression was performed using maximum likelihood estimation of a generalised linear model (positive outcome coded as 1, negative outcome as 0). We employed a logarithmic link function, and heteroscedasticity consistent covariance matrix estimators. [28,31] Statistical significance was defined using a 2-sided significance level of α = 0·05. We also conducted sensitivity analysis on the statistical model using quasi-Poisson and log-binomial regression giving relative risks and logistic regression giving odds ratios.

### Ethics and permissions

Approved by the Health Research Authority Research Ethics Committee (East of England, Essex), REC 20/EE/0157, including use of Section 251 of the 2006 NHS Act authorised by the Confidentiality Advisory Group.

## RESULTS

During the study period reported here, 3297 hospital-admissions with SARS-CoV-2 were identified, resulting in 1938 SARS-CoV-2 positive qualifying hospitalisations: 1190 (27 inferred) with Delta variant and 748 (119 inferred) with Omicron variant. Overall, 142(76 Delta and 66 Omicron) cases were excluded from analysis due to being partially vaccinated, or with an infection acquired in hospital, or both (Figure 1).

**Figure 1:**
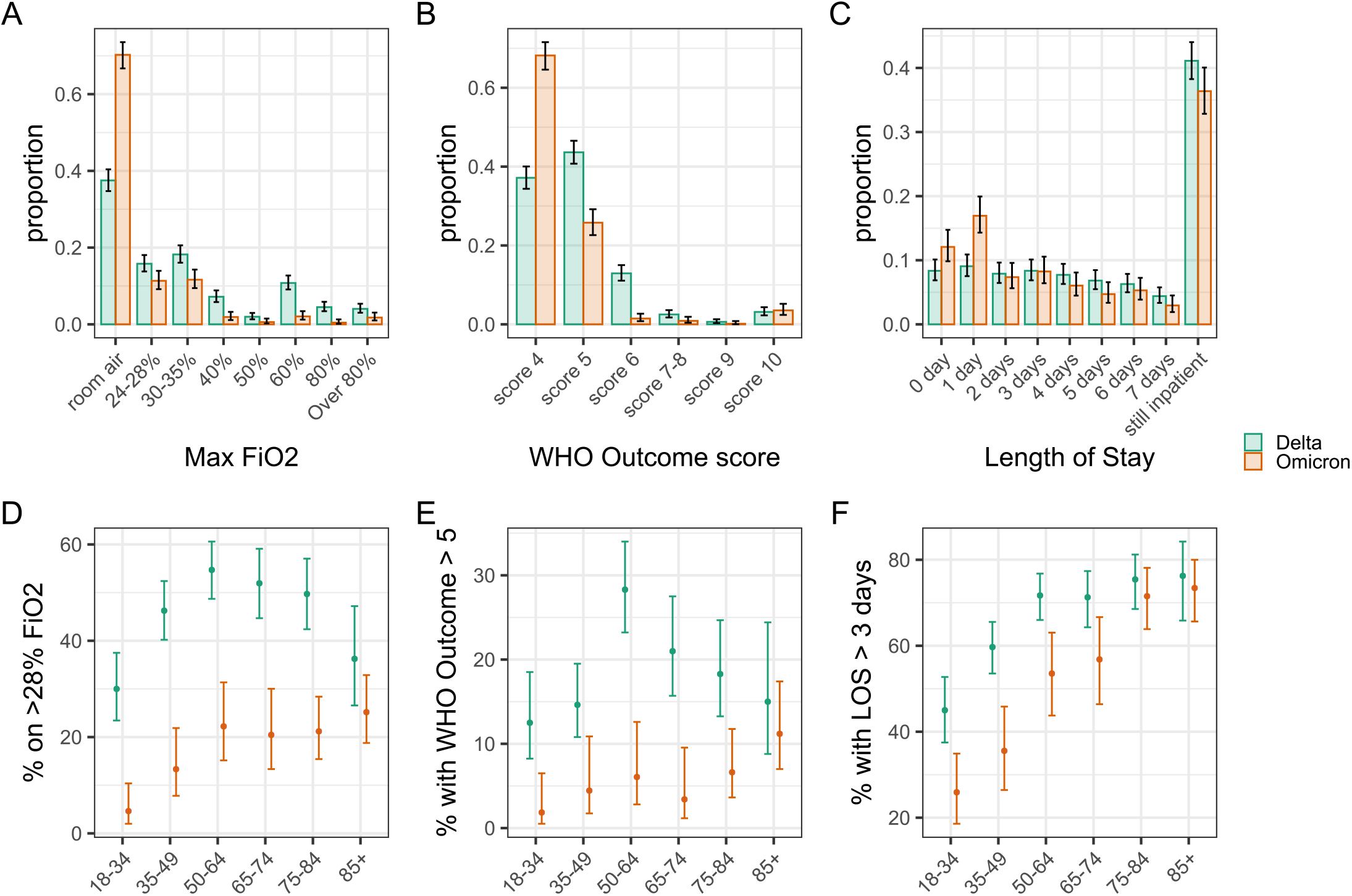
Flow diagram of adults hospitalised with SARS-CoV-2 infection. Inclusion and exclusion criteria in the cohort stratified by the primary comparison or SARS-CoV-2 variant. Any patient may be excluded for multiple reasons at each stage, hence the counts of reasons for exclusion may not add up to the number of patients excluded. FiO_2_, fraction of inspired oxygen; LOS, length of stay; VOC, variant of concern; WHO, World Health Organisation

Patients hospitalised with Omicron variant were statistically significantly older than those with Delta infection (median [IQR]: 70·6 y [44·8y–83·5y] versus 57·7y [42·6y–74·1y] respectively, *P*<0.001) and had a higher CCI (4 [1–5] versus 2 [0–4] respectively, *P*<0·001). These individuals were also more likely to have received a third vaccination dose (60·2% (N=679) versus 5·0% (N=1114) respectively, *P*<0·001) (Table 1). Additional analysis investigated the nature of the additional comorbidities (Supplementary Table 1) and vaccination (Supplementary Table 2), showing that patients admitted with Omicron infection were more likely to have pre-existing heart disease, hypertension, atrial fibrillation, dementia, mild chronic kidney disease (CKD), and peripheral vascular disease than patients admitted with Delta infection (all *P*<0.001).

The proportion of hospitalisations with the Omicron variant increased over time, with Omicron becoming the dominant variant in the last week of December 2021 (Figure 2A). As the vaccination campaign progressed over the study period, we saw an increasing proportion of hospitalised individuals who had received two vaccine doses or booster (Figure 2B). Hospital admissions occurring early in the study tended to be younger: the admission age profile shifted towards older groups with time, with a change point in late December 2021, coincident with increasing Omicron cases (Figure 2C). Overall, the patterns suggest that patients hospitalised between December and February were generally older and frailer, but more commonly vaccinated.

**Figure 2:**
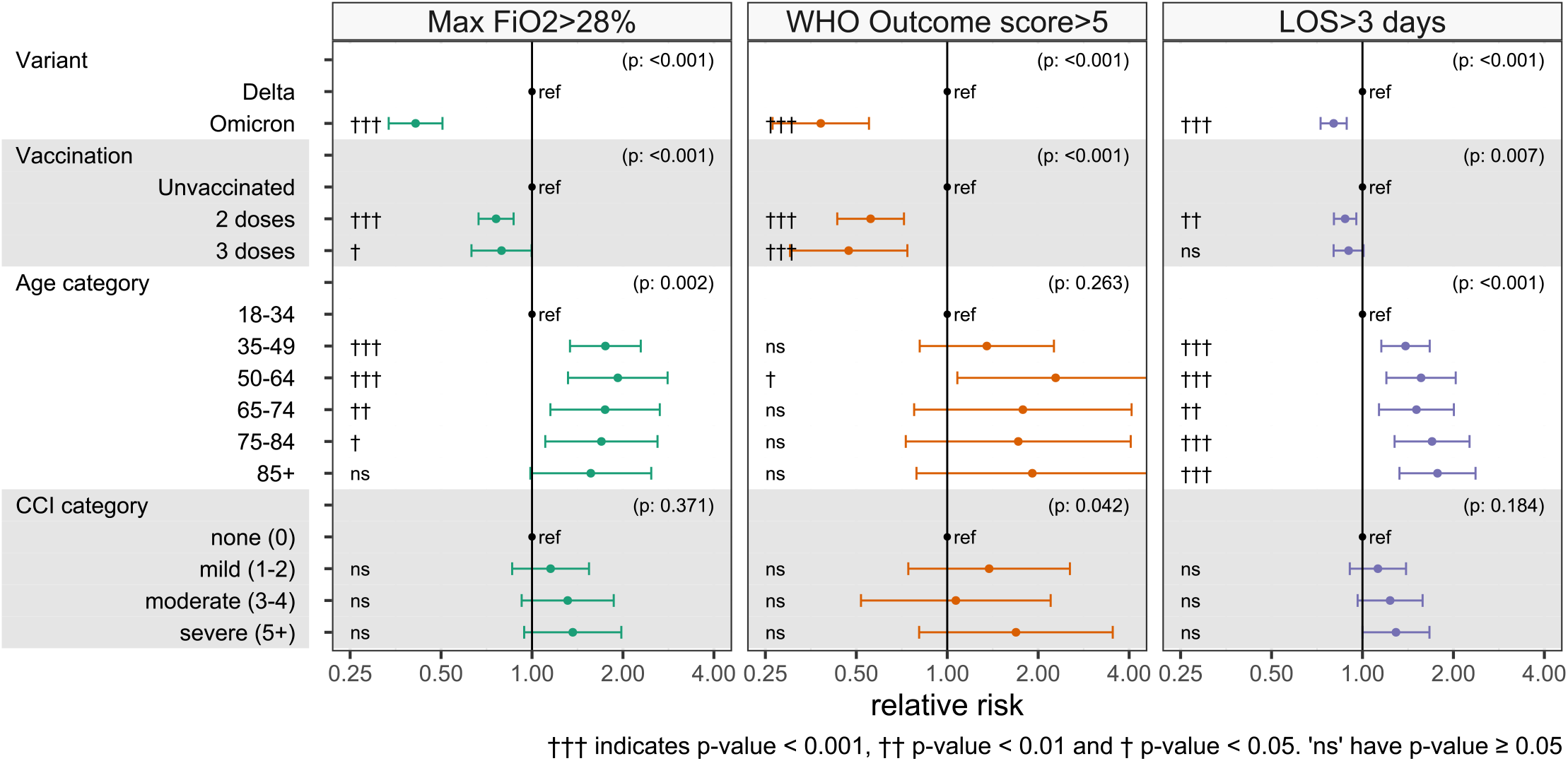
Hospitalisations with SARS-CoV-2 infection throughout the study. (A) Daily rates of hospitalisations with Delta and Omicron SARS-CoV-2 infection, (B) 14-day rolling proportions of hospitalisations by vaccination status (excluding people partially vaccinated on admission) and (C) 14-day rolling proportions of hospitalisations by age groups (in years) throughout the study. Vertical lines represent the earliest time an Omicron case was admitted and the latest time a Delta case was admitted. Before the earliest Omicron detection, cases are assumed to be Delta when sequencing results are not available, and after the last Delta detection, cases are assumed to be Omicron, when sequencing results are not available.

However, patients hospitalised with Omicron infections experienced less severe outcomes compared to patients infected with Delta. Omicron hospitalisations required ventilation less frequently (Figure 3A), and when they did, a lower oxygen supplement was needed (Figure 3B). The WHO outcome score was also lower within the first seven admission days (Figure 3C).

**Figure 3:**
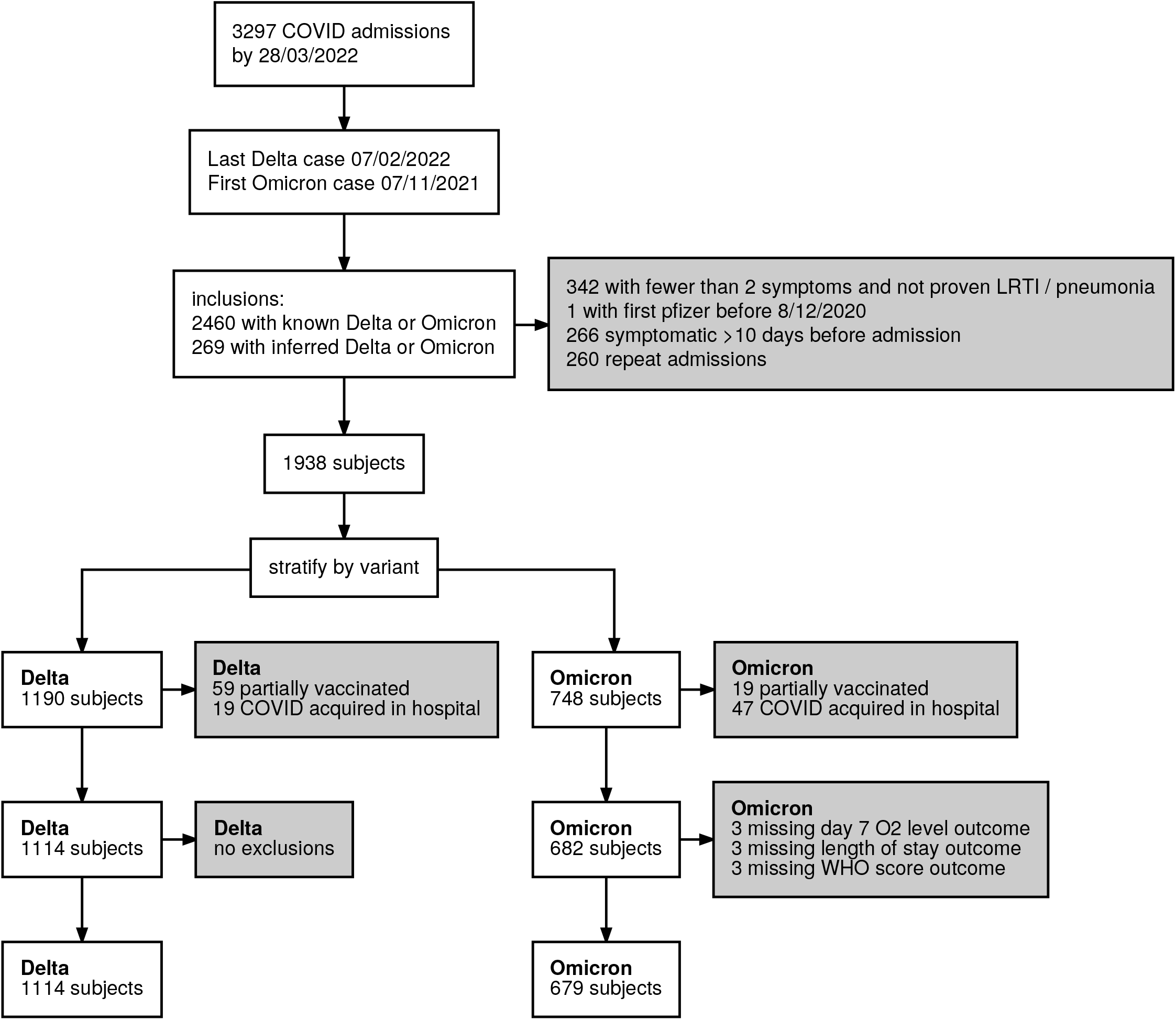
The detailed breakdown of the three selected indicators of severity of hospital admission stratified by SARS-CoV-2 variant, and the comparison of three binary indicators of hospital burden and their relationship to SARS-CoV-2 variant and patient age, for patients admitted to hospital. The distribution of patients (A) requiring different peak levels of oxygen supplementation, (B) with different ventilation requirements as defined by the WHO outcome or clinical progression score, and (C) with different lengths of stay. The proportion of patients who (D) require high flow oxygen >28% FiO_2_, (E) have a WHO outcome score>5 (requiring NIPPV), and (F) have a hospital length of stay greater than three days, as assessed on the seventh day following admission. Error bars show 95% binomial confidence intervals for each outcome, compared to other outcomes. FiO_2_, Fraction of inspired oxygen; LOS, length of stay; NIPPV, non-invasive positive pressure ventilation

There was no discernible difference in these outcomes between Delta and Omicron in patients ≥85 years (Figure 3D, 3E). Length of hospital stay was shorter for Omicron compared to Delta among patients <70 years of age but similar for ≥70 years (Figure 3F).

Direct associations such as unadjusted estimates account for neither the changes in patient frailty nor vaccination status observed over the study period (Figure 3). Figure 4 presents results of adjusted regression models that account for these time-varying factors observed over the study duration (methods in Supplementary Tables 3-11, results in Supplementary Tables 12, 13). Relative to Delta, patients infected with Omicron were less likely to require oxygen supplementation of FiO_2_ >28% [Relative risk (RR)=0·42 (95%CI: 0·34-0·52)], to have a WHO outcome score >5 (which implies that a patient required positive pressure ventilatory support or died in the hospital) [RR=0·33 (95%CI: 0·21-0·50)], and to have a hospital stay lasting over three days [RR=0·84 (95%CI: 0·76-0·92)]. In stratified analyses by vaccination status, infection with Omicron relative to Delta was associated with lower severity across all three measures in both vaccinated and unvaccinated patients.

**Figure 4:**
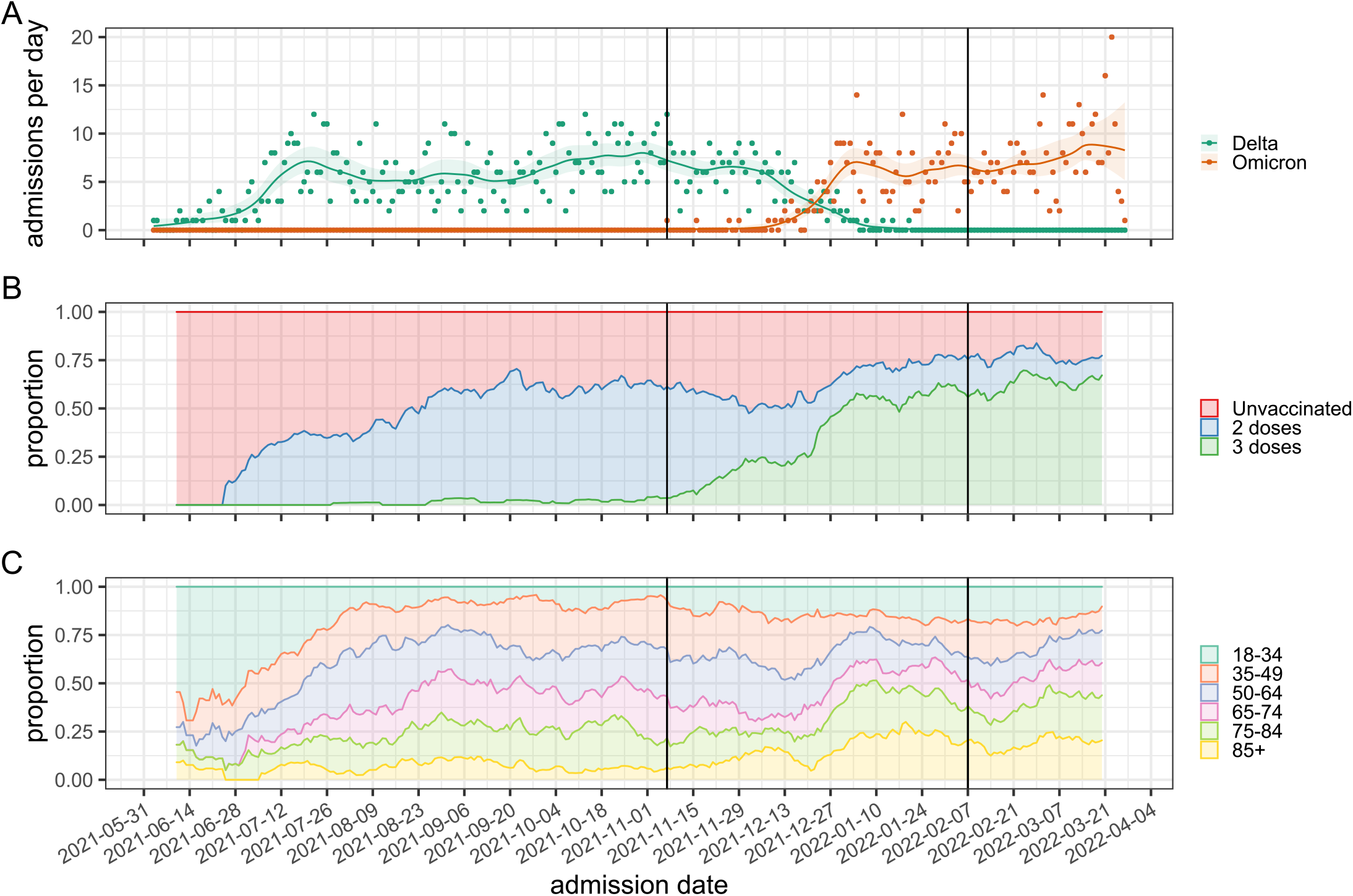
Robust Poisson regression model relative risks for the three indicators of hospital burden. Relative risks describing the effect of different predictors on whether patients require oxygen supplementation with FiO_2_ >28% (first column), have a WHO outcome score greater than 5 (second column) or who remain in hospital for more than 3 days (third column) measured on the seventh day after admission. Numerical details of these regression models, including other explanatory variables that are not our primary interest, are given in Supplementary Table 15. A comparison of relative risks computed by different methods and odds ratios by logistic regression is shown in Supplementary Table 18. FiO_2_ – fraction inspired oxygen, LOS – length of stay, CCI – Charlson Comorbidity Index

Compared to unvaccinated patients, individuals vaccinated with two doses were less likely to require oxygen >28% FiO_2_ [RR=0·78 (95%CI: 0·68-0·89)], positive pressure ventilatory support or increased critical care [RR=0·56 (95%CI: 0·43-0·73)], and to have a hospital admission >3 days [RR=0.90 (95%CI: 0·84-0·98)]. Results for vaccination with three doses were similar but had wider confidence intervals than for two doses. In analyses stratified by infecting variant, vaccination was associated with lower severity of disease using all three measures for both Omicron and Delta infections. In analyses assessing potential effect modification by adding an interaction term between variant and vaccination status to adjusted models, the interaction term was not significant for any severity outcome (Supplementary Table 14).

In sensitivity analyses using threshold levels of increased severity to dichotomize outcomes, the relative lower severity of Omicron declined further when using higher thresholds for oxygen requirement [FiO_2_ >35%: RR=0·27 (95%CI: 0·19-0·39); FiO_2_ >50%: RR=0·23 (95%CI: 0·15-0·37); Supplementary Table 15]. Sensitivity analysis using a WHO outcome score threshold >6 (hence necessitating intubation) had insufficient statistical power (Supplementary Table 16). When longer thresholds for LOS were used, the lower relative severity of Omicron attenuated as the threshold increased [LOS >5 days: RR=0·81 (95%CI: 0·71-0·92); LOS >7 days: RR=0·85 (95%CI: 0·73-0·99); Supplementary Table 17], suggesting certain patient characteristics may be associated with shorter admissions. For vaccination, the protective effect of vaccines increased as the severity level threshold increased for each outcome measure. Associations were similar in sensitivity analyses that replaced age group and CCI with the QCovid2 score.

## DISCUSSION

We found evidence that in adults hospitalised with COVID-19, Omicron infection was associated with less severe illness than that caused by Delta and that vaccination was independently associated with lower in-hospital disease severity using three separate measures of severity. Compared to Delta, Omicron-related hospitalizations were 58%, 67%, and 16% less likely to require high flow oxygen >28% FiO_2_, positive pressure ventilatory support or more critical care, and to have a hospital stay lasting more than three days, respectively. These findings persisted even after controlling for multiple potential confounders including age, chronic medical conditions, frailty, and increasing vaccination coverage over time. Our findings are in keeping with previously published assessment of the relative severity of Omicron variant infection compared to other variants, which examined markers of severity such as hospitalization, ICU admission, mechanical ventilation, severe disease and death [32,33] [11,17,18]. Lauring *et al*. reported lower in-hospital severity for Omicron than for Delta among unvaccinated patients and that vaccinated patients experienced less severe disease regardless of infecting variant[11]. Although studies have reported lower severity for Omicron relative to Delta, measured as the risk of hospitalization [17,18] or death [17] after taking into account vaccination status, the scientific evidence continues to evolve.

However, in determining the reduced requirement of increased oxygen requirement and total positive pressure requirement, including non-invasive ventilation, this analysis should contribute to future hospital care and service planning assessments. The impact of lower severity Omicron-related hospitalization must be balanced with increased transmissibility and overall higher numbers of infections with this variant. Considerable morbidity resulted from Omicron infection, with 18% of Omicron admissions requiring oxygen supplementation FiO_2_ >28%, 6% requiring positive pressure ventilation, 62% needing hospitalization ≥four days, and 4% in-hospital mortality. Despite individual-level risk reduction of needing increasing oxygen supplementation or high dependency care with Omicron compared to earlier variants, healthcare systems could still be overwhelmed by the sheer case numbers and resulting high hospitalization burden, especially if the less severe variant is offset by increasingly frail and elderly patients requiring hospitalization. Omicron’s increased transmissibility also increases the infection risk for clinical and other hospital staff, which can take a significant toll on staffing levels.

The length of hospitalization in individuals who have already developed disease severe enough to merit hospital admission is affected by multiple factors which may confound the effect of any pathogen or strain, including patient frailty and need for continued care following discharge. Other events unrelated to the patient’s primary pathology, such as secondary hospital-acquired infection, may also impact discharge. Therefore, discerning the effect of variant on hospital length of admission is difficult. In contrast, the reductions in oxygen requirements and lower WHO outcome scores we observe for Omicron infections are reassuring. This suggests that exhaustion of oxygen supplies or high dependency care beds are less likely in large waves of community Omicron infection than with Delta and other previous variants. Vaccination reduces these risks further and our results suggest it is an important modifier of the impact of these aspects on health services.

Our study has several strengths. All patients were admitted to hospital with acute respiratory illness caused by SARS-CoV-2, so these results are unlikely to be subject to bias caused by admission for other causes (i.e., incidental COVID-19 disease). In the UK, all national programme vaccines are provided at no cost at the point of delivery; hence our cohort is not biased by issues surrounding the cost of vaccination that are seen in fee-based systems [34]. While week of symptom onset was identified as a significant univariate predictor for adverse outcome, we observe it becomes non-significant on a multivariable analysis which includes time varying factors, such as vaccination, or variant status (Supplementary Tables 6, 8, 10). This reassures us that there are no other time varying factors for which we have not accounted. With a comparatively rich data set of prospectively collected information, we can determine timing of disease onset accurately and capture information on a range of factors more precisely, such as vaccination status and patient comorbidities, than could be done using a retrospective design.

However, we are limited in this study to observation of patient outcomes in patients only after hospital admission. Thus, we cannot draw conclusions about the severity of Omicron infections occurring entirely in the community. We observed a change in the demographics of patients being admitted towards more elderly and frail populations, which could be explained if community Omicron disease were especially milder in young and healthy individuals. The effects we observed for Omicron and vaccination on clinical outcomes in hospitalised patients occurred over and above any community-level effects. In this study, lineage association was only done using lineage-specific PCRs, and it was not possible to attribute a particular case to an exact sub-lineage. During the period of this analysis the Omicron sub-lineages circulating in the UK were BA.1 and to a small extent BA.2. The Delta sub-lineages were principally undifferentiated B.1.617.2 and AY.4, as shown in Supplementary Figure 1. Estimates presented here are for the combined effect of the sub-lineages. We did not have sufficient data to determine the effectiveness of any individual vaccine against the outcomes studied and our results cannot be interpreted as a vaccine effectiveness estimate. Furthermore, we did not have sufficient statistical power to determine with confidence whether booster doses of COVID-19 vaccine offered additional protection against adverse outcomes. We are unable to control for pressures in the social care system following discharge, and large waves of community Omicron infection may put pressure on community elderly care facilities resulting in a backward pressure on hospital discharges. This could partially explain our observation that length of stay longer than three days is less associated with variant status and vaccination than other outcomes. Although dexamethasone was used as treatment in both Delta and Omicron waves, novel antivirals were introduced throughout the study and this may affect results. Whilst the population of the Bristol area is representative of the UK population, it is principally Caucasian, and our findings must be interpreted in this context. Routine testing for anti-nucleocapsid antibodies was not conducted, and we therefore do not have these data available for determining previous infection status of hospitalised individuals within the Bristol area nor how our findings were impacted by population-level changes in infection-derived or hybrid immunity over time. The study design does not involve any clustering of observations, but we cannot exclude spontaneous formation of clusters in the data set due to outbreaks in the catchment area, which we can only partly mitigate by including covariates of age, CCI and vaccination status.

In this prospective study, patients hospitalised with Omicron infections had lower clinical severity than those with Delta infections, as assessed by maximal oxygen requirements, a validated WHO outcome score, and hospital length of stay. Omicron infection, however, still resulted in substantial patient and public health burden following admission. Older and frail patients remain particularly susceptible to severe SARS-CoV-2 infection, underscoring the importance of maintaining high levels of vaccine coverage in this population. Understanding differences in the risk of clinical disease caused by SARS-CoV-2 variants following infection and following hospitalisation, and the role of vaccination and other factors in modifying risk, is critical for planning of public health measures.

## Data Availability

The data used in this study are sensitive and cannot be made publicly available without breaching patient confidentiality rules.

## CONTRIBUTORS

CH, RC, RM, LD, JO, JN, JM and AF generated the research questions and analysis plan. CH, AM, JK, MC and The Avon CAP team were involved in data collection. CH, RC, RM, LD, and AF undertook data analysis. All authors (CH, RC, RM, JN, EB, JS, JK, AM, JK, MC, JO, GE, NM, LJ, SG, BG, JMM, LD and AF) were involved in the final manuscript preparation and its revisions before publication. AF provided oversight of the research.

## DATA SHARING STATEMENT

The data used in this study are sensitive and cannot be made publicly available without breaching patient confidentiality rules. All analysis code is available on GitHub: https://doi.org/10.5281/zenodo.7220569

## ACKNOWLEDGEMENTS

The authors would like to thank the UK Health Security Agency (UKHSA) Vaccine Effectiveness Working group and the University of Bristol UNCOVER group for guidance in data analysis and study design. We thank colleagues at the University of Bristol for their support with this study, including Rachel Davies, Paul Savage, Emma Foose, Susan Christie, Mark Mummé, and Adam Taylor. We want to recognise the help from Kevin Sweetland and Aman Kaur-Singh in the AvonCAP study. We would also like to acknowledge the research teams at North Bristol and University Hospitals of Bristol and Weston NHS Trusts for making this study possible, including Helen Lewis-White, Rebecca Smith, Rajeka Lazarus, Mark Lyttle, Kelly Turner, Jane Blazeby, Diana Benton, and David Wynick. We would also like to acknowledge all participants of the many studies undertaken to find effective vaccines against SARS-CoV-2.

## The AvonCAP Research Group

Anna Morley, Amelia Langdon, Anabella Turner, Anya Mattocks, Bethany Osborne, Charli Grimes, Claire Mitchell, David Adegbite, Emma Bridgeman, Emma Scott, Fiona Perkins, Francesca Bayley, Gabriella Ruffino, Gabriella Valentine, Grace Tilzey, James Campling, Johanna Kellett Wright, Julia Brzezinska, Julie Cloake, Katarina Milutinovic, Kate Helliker, Katie Maughan, Kazminder Fox, Konstantina Minou, Lana Ward, Leah Fleming, Leigh Morrison, Lily Smart, Louise Wright, Lucy Grimwood, Maddalena Bellavia, Madeleine Clout, Marianne Vasquez, Maria Garcia Gonzalez, Milo Jeenes-Flanagan, Natalie Chang, Niall Grace, Nicola Manning, Oliver Griffiths, Pip Croxford, Peter Sequenza, Rajeka Lazarus, Rhian Walters, Robin Marlow, Robyn Heath, Rupert Antico, Sandi Nammuni Arachchge, Seevakumar Suppiah, Taslima Mona, Tawassal Riaz, Vicki Mackay, Zandile Maseko, Zoe Taylor, Zsolt Friedrich, Zsuzsa Szasz-Benczur.

## REFERENCES

1. Challen R, Dyson L, Overton CE, et al. Early epidemiological signatures of novel SARS-CoV-2 variants: establishment of B.1.617.2 in England. medRxiv 2021: 2021.06.05.21258365.

2. Government U. Coronavirus (COVID-19) in the UK. 2022. https://coronavirus.data.gov.uk/details/vaccinations?areaType=overview&areaName=United%20Kingdom (accessed 14 February 2022).

3. UKHSA. SARS-CoV-2 variants of concern and variants under investigation in England, Technical briefing 36. 2022.

4. Wang P, Nair MS, Liu L, et al. Antibody resistance of SARS-CoV-2 variants B. 1.351 and B. 1.1. Nature 2021; 593(7857): 130–5.

5. Liu Y, Liu J, Xia H, et al. Neutralizing activity of BNT162b2-elicited serum. New England Journal of Medicine 2021; 384(15): 1466–8.

6. Nemet I, Kliker L, Lustig Y, et al. Third BNT162b2 Vaccination Neutralization of SARS-CoV-2 Omicron Infection. New England Journal of Medicine 2021; 386(5): 492–4.

7. Schmidt F, Muecksch F, Weisblum Y, et al. Plasma Neutralization of the SARS-CoV-2 Omicron Variant. New England Journal of Medicine 2021; 386(6): 599–601.

8. Ferguson N, Ghani A, Hinsley W, Volz E. Report 50: Hospitalisation risk for Omicron cases in England. Imperial College London 2022.

9. Sheikh A, Kerr S, Woolhouse M, McMenamin J, Robertson C. Severity of Omicron variant of concern and vaccine effectiveness against symptomatic disease: national cohort with nested test negative design study in Scotland. University of Edinburgh 2021.

10. UKHSA. COVID-19 vaccine surveillance report: Week 11: UK Government, 2022.

11. Lauring AS, Tenforde MW, Chappell JD, et al. Clinical severity of, and effectiveness of mRNA vaccines against, covid-19 from omicron, delta, and alpha SARS-CoV-2 variants in the United States: prospective observational study. BMJ 2022; 376: e069761.

12. Ferdinands J, Rao S, Dixon B, et al. Waning 2-Dose and 3-Dose Effectiveness of mRNA Vaccines Against COVID-19–Associated Emergency Department and Urgent Care Encounters and Hospitalizations Among Adults During Periods of Delta and Omicron Variant Predominance — VISION Network, 10 States, August 2021–January 2022. MMWR Morb Mortal Wkly Rep 2022; 71: 255–63.

13. Thompson M, Natarajan K, Sa I, et al. Effectiveness of a Third Dose of mRNA Vaccines Against COVID-19–Associated Emergency Department and Urgent Care Encounters and Hospitalizations Among Adults During Periods of Delta and Omicron Variant Predominance — VISION Network, 10 States, August 2021–January 2022. MMWR Morb Mortal Wkly Rep 2022; 71: 139–45.

14. Jassat W, Abdool Karim SS, Mudara C, et al. Clinical severity of COVID-19 in patients admitted to hospital during the omicron wave in South Africa: a retrospective observational study. The Lancet Global Health.

15. Kirsebom FCM, Andrews N, Stowe J, et al. COVID-19 vaccine effectiveness against the omicron (BA.2) variant in England. Lancet Infect Dis 2022.

16. UKHSA UHSA. COVID-19 vaccine surveillance report: week 19. 2022. https://assets.publishing.service.gov.uk/government/uploads/system/uploads/attachment_data/file/1075115/COVID-19_vaccine_surveillance_report_12_May_2022_week_19.pdf (accessed 8th June 2022 2022).

17. Nyberg T, Ferguson NM, Nash SG, et al. Comparative analysis of the risks of hospitalisation and death associated with SARS-CoV-2 omicron (B.1.1.529) and delta (B.1.617.2) variants in England: a cohort study. The Lancet.

18. Bager P, Wohlfahrt J, Fonager J, et al. Risk of hospitalisation associated with infection with SARS-CoV-2 lineage B.1.1.7 in Denmark: an observational cohort study. Lancet Infect Dis 2021; 21(11): 1507–17.

19. Strasser Z, Hadavand A, Murphy S, Estiri H. SARS-CoV-2 Omicron Variant is as Deadly as Previous Waves After Adjusting for Vaccinations, Demographics, and Comorbidities. Research Square 2022.

20. Hyams C, Marlow R, Maseko Z, et al. Effectiveness of BNT162b2 and ChAdOx1 nCoV-19 COVID-19 vaccination at preventing hospitalisations in people aged at least 80 years: a test-negative, case-control study. The Lancet Infectious Diseases 2021; 21(11): 1539–48.

21. Harris PA, Taylor R, Thielke R, Payne J, Gonzalez N, Conde JG. Research electronic data capture (REDCap)—A metadata-driven methodology and workflow process for providing translational research informatics support. Journal of Biomedical Informatics 2009; 42(2): 377–81.

22. Charlson ME, Pompei P, Ales KL, MacKenzie CR. A new method of classifying prognostic comorbidity in longitudinal studies: Development and validation. Journal of Chronic Diseases 1987; 40(5): 373–83.

23. Hippisley-Cox J, Coupland CAC, Mehta N, et al. Risk prediction of covid-19 related death and hospital admission in adults after covid-19 vaccination: national prospective cohort study. BMJ 2021; 374:n2244.

24. Rockwood K, Song X, MacKnight C, et al. A global clinical measure of fitness and frailty in elderly people. CMAJ : Canadian Medical Association journal = journal de l’Association medicale canadienne 2005; 173(5): 489–95.

25. Physicians RCo. National Early Warning Score (NEWS) 2: Standardising the assessment of acute-illness severity in the NHS. Updated report of a working party. RCP, London 2017.

26. UKHSA. SARS-CoV-2 variants of concern and variants under investigation in England, Technical Briefing 33, 2021.

27. Characterisation WHOWGotC, Management of C-i. A minimal common outcome measure set for COVID-19 clinical research. Lancet Infect Dis 2020; 20(8): e192–e7.

28. Zou G. A modified poisson regression approach to prospective studies with binary data. Am J Epidemiol 2004; 159(7): 702–6.

29. van Buuren S, Groothuis-Oudshoorn K. mice: Multivariate Imputation by Chained Equations in R. Journal of Statistical Software 2011; 45(3): 1–67.

30. Team RC. R: A language and environment for statistical computing.. In: Computing RFfS, editor. Vienna, Austria; 2014.

31. Zeileis A, Köll S, Graham N. Various Versatile Variances: An Object-Oriented Implementation of Clustered Covariances in R. Journal of Statistical Software 2020; 95(1): 1–36.

32. Lewnard JA, Hong VX, Patel MM, Kahn R, Lipsitch M, Tartof SY. Clinical outcomes associated with SARS-CoV-2 Omicron (B.1.1.529) variant and BA.1/BA.1.1 or BA.2 subvariant infection in southern California. Nature Medicine 2022.

33. Van Goethem N, Chung PY, Meurisse M, et al. Clinical Severity of SARS-CoV-2 Omicron Variant Compared with Delta among Hospitalized COVID-19 Patients in Belgium during Autumn and Winter Season 2021–2022. Viruses 2022; 14(6).

34. Wheelock A, Miraldo M, Thomson A, Vincent C, Sevdalis N. Evaluating the importance of policy amenable factors in explaining influenza vaccination: a cross-sectional multinational study. BMJ Open 2017; 7(7): e014668.

