## supplementary data 1 for "Severity of Omicron (B.1.1.529) and Delta (B.1.617.2) SARS-CoV-2 infection among hospitalised adults: a prospective cohort study in Bristol, United Kingdom"

**UK Vaccination Programme**

The UK vaccine programme prioritised first dose administration, extending the interval between first and second doses of COVID-19 vaccines up to 12 weeks [1]. Initially BNT162b2 (Comirnaty®) was deployed in the UK, followed rapidly by the addition of ChAdOx1 (Vaxzevria®) and, subsequently, mRNA-1273 (Spikevax®). After January 2022, coverage with 2 doses of vaccine was over 80% in adults aged ≥20 years of age with higher rates among those ≥50 years of age [2]. A booster campaign delivering a third vaccine dose commenced in September 2021 and was accelerated in December 2021 to meet the emerging threat of Omicron [3], resulting in 60% of adults receiving a booster by 1st January 2022. Both the primary vaccination campaign and the booster campaign were rolled out in an age-stratified fashion, with elderly and high-risk groups targeted first [4].

**References for Supplementary Data 1:**

1.	JCVI. Optimising the COVID-19 vaccination programme for maximum short-term impact. 26th January 2021 2021. <https://www.gov.uk/government/publications/prioritising-the-first-covid-19-vaccine-dose-jcvi-statement/optimising-the-covid-19-vaccination-programme-for-maximum-short-term-impact> (accessed 24th February 2021).

2.	UKHSA. COVID-19 vaccine surveillance report - Week 51. 2021.

3.	England N. NHS begins COVID-19 booster vaccination campaign. 2021.

4.	Government U. Coronavirus (COVID-19) in the UK. 2022. <https://coronavirus.data.gov.uk/details/vaccinations?areaType=overview&areaName=United%20Kingdom> (accessed 14 February 2022).

**Supplementary Figure 1:**

**Variant proportions**

The proportions of different genomic variants of SARS-CoV-2 identified over time by COVID-19 Genomics UK Consortium (COG-UK) as made available by the Sanger centre (<https://covid19.sanger.ac.uk/lineages/raw>). Raw weekly case counts for all lineages are combined with the Pango lineage hierarchy files (<https://github.com/cov-lineages/pango-designation>) and fed into a multinomial log-linear regression model optimised with a single hidden layer neural network using the R package ‘nnet’ (<https://rdocumentation.org/packages/nnet/versions/7.3-17>, citation here: <https://cran.r-project.org/web/packages/nnet/citation.html> ). The study period is marked by vertical lines labelled “start” and “end”. During the study variants in general circulation included the Delta lineages B.1.617.2, and AY.4 (orange), and the Omicron lineages BA.1 and BA.2 (blue). Other significant dates marked by vertical lines are the 7^th^ of November, before which all infections are assumed to be Delta variant, and the 7^th^ of February after which all infections are assumed to be Omicron. In between these dates lineage designation was only done following explicit testing using lineage specific PCR tests. These tests only differentiated Delta from Omicron and did not give information about the sub-lineages.

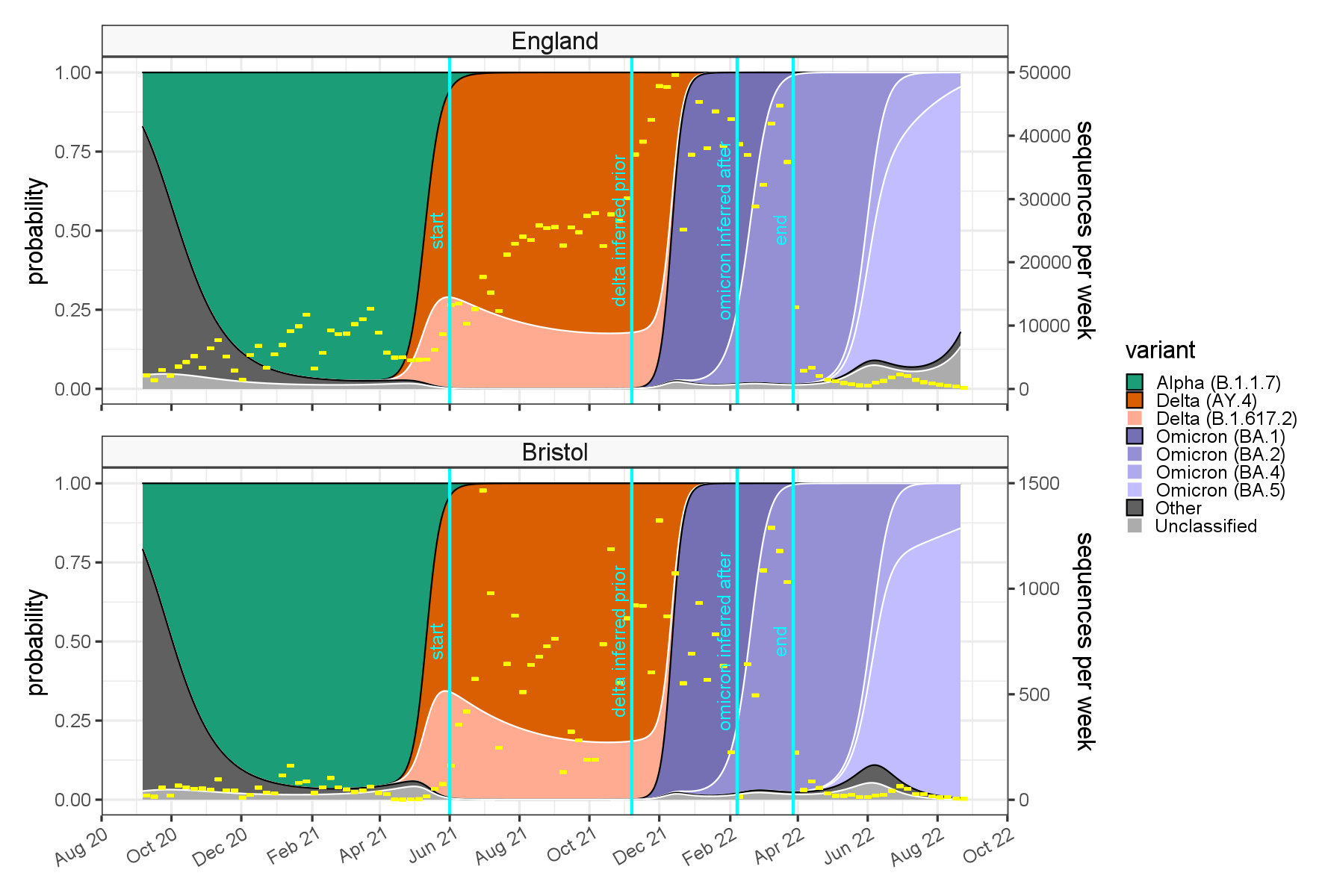
